# Demographic Drivers of Epidemic Outcomes: Sensitivity Analysis of Multidimensional Parameters in the Covasim Model

**DOI:** 10.1101/2025.10.14.25328629

**Authors:** Vera Tsurkis, Ivan Kozlov, Andrei Samoilov, Irina Maslova, Elena Ilina, Alexander Lukashev, Alexander Manolov

**Author notes:** Corresponding author. Ludwig-Maximilian University of Munich, Faculty of Physics; Geschwister-Scholl-Platz 1, D-80539 Muenchen.

## Abstract

**Background:** Sensitivity analysis is a key tool for identifying which model inputs most strongly influence model outputs thereby informing data collection priorities. In agent-based models, these inputs include demographic parameters used to construct synthetic populations. Such parameters — age distributions, household size distributions, and contact matrices — are critical in shaping transmission dynamics. However, most established sensitivity analysis methods are designed for scalar inputs and cannot readily accommodate multidimensional demographic data. We introduce a novel sampling approach to assess the influence of such parameters on model outputs, addressing a key gap in sensitivity analysis of epidemiological agent-based models.

**Methods:** The Covasim model of COVID-19 transmission was used as the case study in this work. An autoencoder neural network was trained on country-level demographic datasets to generate realistic samples of high-dimensional inputs of the model. These sampled inputs were coupled with Sobol’ sensitivity indices to quantify their influence on cumulative infections and deaths.

**Results:** Non-scalar parameters were found to exert a major influence on model outputs. Household size distribution was the most important parameter for cumulative number of infectious cases, while age distribution had the largest effect on cumulative deaths. These findings were consistent across experimental settings, and parameter rankings were stable despite stochastic variation.

**Conclusions:** Our autoencoder-based sampling approach extends methods of global sensitivity analysis to high-dimensional demographic parameters in agent-based epidemiological models. Our results highlight that only a subset of demographic inputs exert a dominant influence on Covasim outputs, and similar behavior can be expected in other epidemiological modeling platforms.

## Introduction

Coronavirus pandemic highlighted the need for detailed models which could be used for reliable forecasts and evaluation of interventions. To obtain a realistic model of an outbreak, one has to use appropriate synthetic population.

The synthetic populations of the epidemiological agent-based models are often constructed using national population censuses as a major source of information. For example, agent-based model was used to simulate the spread of measles in Ireland town in 2012; the census data from Ireland’s Central Statistics office was the main source of information [1]. Additionally, transportation surveys may be included to produce a realistic structure of social contact networks: e.g. USA National Household Transportation Survey was used in EpiSindemics [2] and the Framework for Reconstructing Epidemic Dynamics (FRED) [3]. Cell phone tracking can be also used to obtain realistic individual movement patterns [4]. A major challenge for modelers is to obtain the data that is both sufficiently precise and up-to-date: unfortunately, censuses are not frequent enough to provide recent information about the current population, and transportation or mobile tracking data are not always available. This raises the question of whether all model parameters require such detailed data. Indeed, if a given parameter does not significantly influence the model outcome, it can be roughly estimated or excluded from the model for simplification. Usually, the importance of the parameters is determined through the sensitivity analysis [6].

Sensitivity analysis is a procedure which matches uncertainty of the model’s output with uncertainties in input parameters, therefore, it shows to what extent each parameter influences the outcome [6].

There are two types of sensitivity analysis: local and global [7]. Local sensitivity analysis refers to the analysis of the model’s behavior in close proximity of a particular point in the parametric space (e.g. default values) [8]. The most straightforward method of local analysis is the ‘one-at-a-time’ method, when all but one parameter are fixed and this one parameter is varied around its initial value [9]. It should be noted that the results of local analysis depend on the choice of the point around which the analysis is performed [10]. In contrast, global analysis explores a large part of the parametric space of the model [11]. Among others, the variance-based methods of the global sensitivity analysis are widely used, such as Sobol’ sensitivity analysis [12] and Fourier amplitude sensitivity test (FAST) [13] Due to the ease of interpretation of their results, these methods are often favored over alternative approaches, such as ensemble learning models (random forests), which can also be applied to sensitivity analysis in similar contexts [14].

Sobol’ indices in particularly are frequently used for global sensitivity analysis of agent-based models and epidemiological analytical models. Their application is straightforward and doesn’t rely on analytical calculations which are impossible to apply to agent based models (e.g., partial derivatives). For example, Sobol’ method of sensitivity analysis was applied to the Soil and Water Assessment Tool (SWAT) model, to determine the most important parameters and pairs of parameters [15]. Similarly, Sobol’ indices were used to identify the most important parameters of the agricultural land conservation agent-based model and to subsequently reduce input space by setting non-influential parameters to default values [16]. In epidemiological studies Sobol’ indices were applied to the generalized SIR model of the COVID-19 spread [17].

In the majority of studies Sobol’ method is applied to scalar parameters. However, many population-level parameters are represented as vectors or matrices that possess intrinsic structure. The elements of these vectors and matrices are not independent and therefore cannot be sampled independently. One way to assess sensitivity with respect to non-scalar parameters is to use a low-dimensional space generated by an autoencoder.

Autoencoders are neural network architectures designed for unsupervised learning. They have been theoretically introduced in 1986 [18], and with the growth of computer power became a robust tool for many scientific areas. Autoencoders are used in many fields of biology, especially in bioinformatics: in genomics [19], transcriptomics [20], proteomics [21]. They are trained by minimizing the difference (parameterized by some metric) between the initial and reproduced data. In this process an autoencoder learns compressed representations of data by encoding inputs into a lower-dimensional latent space and then decoding them back to reconstruct the original input. Once trained, the latent space of an autoencoder constitutes an effective low-dimensional representation of the input data and can be used for sampling of synthetic data through the sampling of points in the latent space and further decoding of these points. In this work we apply Sobol’ method coupled with sampling of vector and matrix parameters via autoencoders to the Covasim epidemiological model [22].

Since its release in 2021 the Covasim model has become one of the most widely used agent-based models for studying the spread of SARS-CoV-2 and other respiratory viruses [22] [23] [24] [25]. Here, we assess sensitivity of the Covasim model to demographic parameters of a synthetic population.

## Materials and data

The Covasim model is considered in this paper. It is described in detail in [22]. Shortly, the virus is transmitted between agents via their contacts. All contacts are divided into 4 layers: household, school, work, and community contacts. Probability of transmission depends on the agent’s age.

The core part of the experiments consisted of multiple runs of Covasim with different parameter sets. There were two steps in each run: first, a synthetic population was made (using Synthpops [26]); then, the spread of virus in this population was modeled. For all experiments the Covasim version 3.1.6 (cloned from github at https://github.com/InstituteforDiseaseModeling/covasim) and Synthpops version 1.10.5 (cloned from github at https://github.com/InstituteforDiseaseModeling/synthpops) were used.

## Sampling method

17 demographic parameters were included in the analysis: all demographic parameters in Synthpops and the average number of community contacts in Covasim, as this layer is not constructed in Synthpops. All parameters are listed in Table 1.

**Table 1.** Information about parameters under consideration.

| Parameter | Short name | Sampling method | Sampling range | Data source |
| --- | --- | --- | --- | --- |
| Contact matrices | {layer}_conts | Autoencoder with 1D latent space | [5 percentile, 95 percentile] in latent space | Synthetic contact matrices from [30] |
| Age distribution | age_distr | Autoencoder with 2D latent space | Rectangle in latent space | World Population Prospects 2024 [31] |
| Household size distribution | hh_size | Autoencoder with 1D latent space | [5 percentile, 95 percentile] in latent space | CORESIDENCE Database (CoDB) [32] |
| Household head age by household size | hha_bysize | Autoencoder with 1D latent space | [5 percentile, 95 percentile] in latent space | Demographic Statistics Database United Nations Statistics Division [33] |
| Enrolment rate | enrolment | Range from statistical data | [5 percentile, 95 percentile] in each bin | OECD Education at a Glance [34] |
| Employment rate | employment | Autoencoder with 1D latent space | [5 percentile, 95 percentile] in latent space | International Labour Organization (ILOSTAT) [35] |
| Workplace size distribution | work_distr | Range from statistical data | [5 percentile, 95 percentile] in each bin | Eurostat [36] |
| School size distribution | school_distr | Multiplication by coefficient | [0.5, 2] | — |
| Average class size | class_size | Range from statistical data | [5 percentile, 95 percentile] ([14, 30]) | OECD Education at a Glance [34] |
| Average student teacher ratio | stud_teach | Range from statistical data | [5 percentile, 95 percentile] ([6.3, 27.2]) | OECD Education at a Glance [34] |
| Average teacher teacher degree (average number of contacts between teachers per person per day) | teach_deg | Multiplication by coefficient | [0.5, 2] | — |
| The average number of students per staff members at school | stud_all | Multiplication of the average student teacher ratio by coefficient | [0.5, 1] | — |

Table 1 – *Continued from previous page*
| Parameter | Short name | Sampling method | Sampling range | Data source |
| --- | --- | --- | --- | --- |
| Average additional staff degree (average number of contacts for non-teaching staff member per person per day) | staff_deg | Multiplication by coefficient | [0.5, 2] | — |
| Average number of community contacts (per person per day) | cv_conts | Range from statistical data | [5 percentile, Covasim’s default value] ([4, 20]) | Synthetic contact matrices from [30] + Age distribution [31] |

To obtain comprehensive coverage of the parameter space using the Sobol’ sequence, it is necessary to define the bounds within which the parameters vary. Whenever possible, real-world statistical data from different countries were utilized to constrain parameters and avoid unrealistic values. The set of countries used for such evaluations varied by parameter, depending on data availability. In general, data from Organisation for Economic Co-operation and Development (OECD) countries were used and supplemented with data from other regions when available. African countries were excluded due to variation in population parameters, which often rendered them incompatible with non-African countries and led to failures in constructing synthetic populations using SynthPops.

The overall logic of the experimental workflow is illustrated in Fig. 1A. When sufficient statistical data were available, most parameters represented as matrices or distributions were sampled using autoencoders. This neural network architecture was selected due to the convenience of using its latent space to generate new, realistic parameter values.

**Fig 1.**
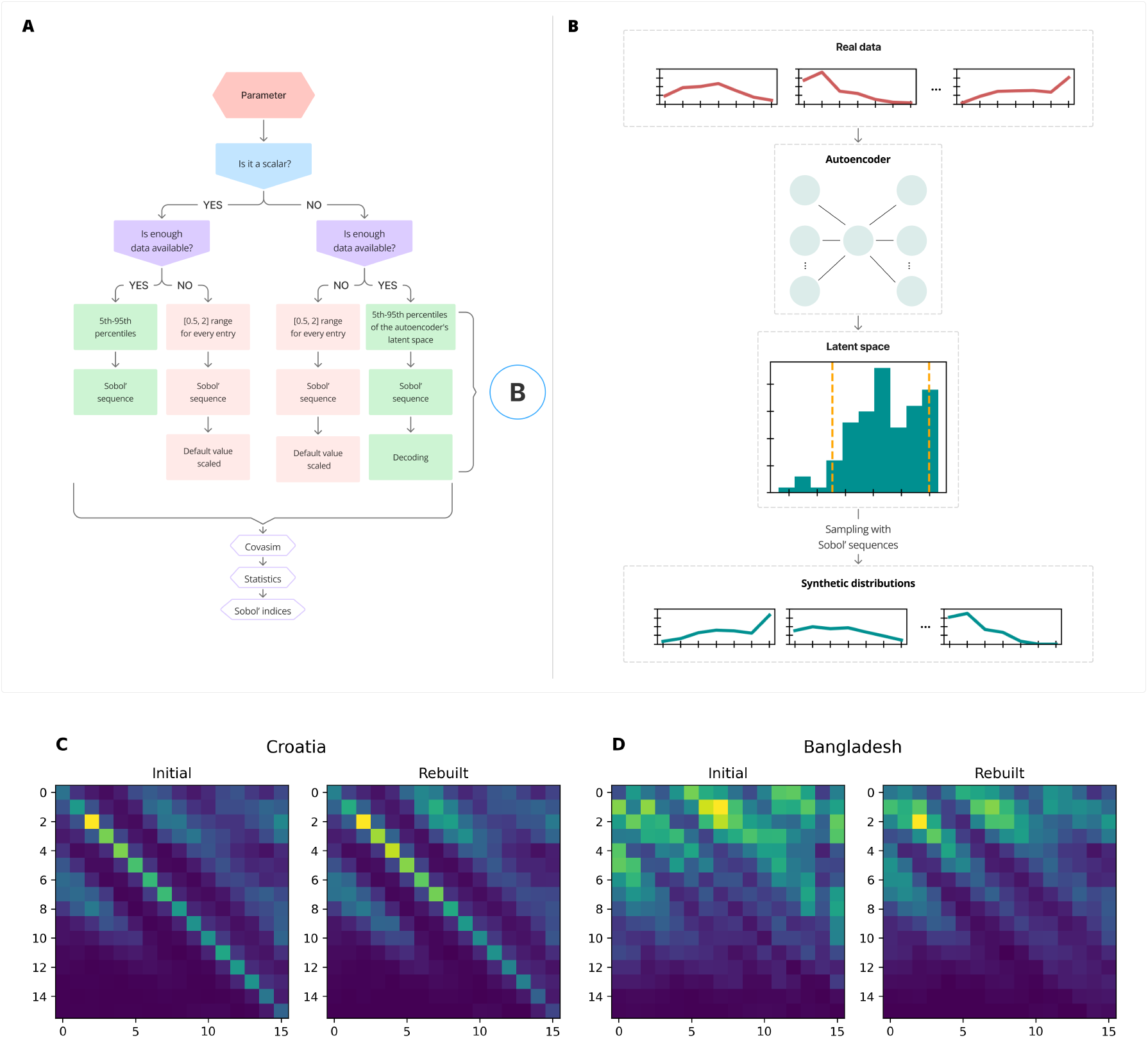
The overall pipeline of the experiments. A – sampling of parameters depending on their dimensionality and the availability of statistical data; B – special case of sampling using autoencoders for matrix- or distribution-based parameters; C, D - examples of initially provided to autoencoder matrices and recovered once

Details of the autoencoder training procedure are presented in Fig. 1B.

All autoencoders trained in this study were based on the same architecture: a small (1-3) number of linear layers connecting the input to the latent space, and a symmetric set of layers mapping from the latent space to the output. While all autoencoders were initially designed with a one-dimensional latent space, this configuration did not yield satisfactory reconstruction results for the age distribution of the population, and a two-dimensional latent space had to be used instead. It should be noted that all autoencoders were initially trained on the full dataset, including African countries where applicable. However, the parameter variation ranges were inferred solely from non-African countries. The performance of the autoencoders is demonstrated in Fig. 1C and Fig. 1D, using the case of household contact matrix sampling as an example. Plots of learning curves for all autoencoders can be found in Supplementary file 1. Although the reconstruction ability of trained autoencoders varied depending on the demographic parameter, each model enabled realistic sampling of the respective parameter from the latent space.

For scalar parameters, realistic values were estimated from statistical data where available, and the interval between the 5th and 95th percentiles was selected to define the range of variation. For workplace size distribution and enrollment rate this procedure was applied to every bin of the distribution, since real data were available, but insufficient for reliable autoencoder training. The upper limit for the average number of community contacts was set to the Covasim’s default value, which exceeds the 95th percentile of real data. For some parameters, no publicly available statistical data could be found. For such parameters, the default value was multiplied by a coefficient sampled from the range [0.5, 2]. For school size distribution the same technique was used within every bin of distribution.

A detailed description of the data sources and the ranges chosen can be found in Table 1.

Other simulation parameters were fixed as follows: the population size was fixed at 100,000 individuals and the initial number of infected individuals was set at 30. To account for stochastic effects, each parameter set was simulated using different random seed.

## Sensitivity analysis method

Sensitivity analysis was conducted using Sobol’ indices [12], a method based on functional ANOVA decomposition of the model variance [27].

Let us consider a model *Y* = *f* (*X*_1_, …, *X*_*k*_), where *X*_1_, …, *X*_*k*_ are parameters of the model, Y is a scalar output of the model. Then the first order Sobol’ indices are defined as

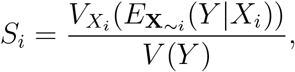

where *E*_**X**_~*i* (*Y X*_*i*_) is the mathematical expectation of model’s output when parameter *X*_*i*_ is fixed, and the numerator is the variance of this expectation, calculated over all values of *X*_*i*_. The denominator is the variance of the model calculated over the whole parametric space. Similarly, the total Sobol’ indices are defined as

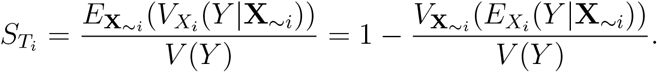

Conceptually, the first-order index *S*_*i*_ represents the fraction of the model’s variance that would be eliminated if the parameter *X*_*i*_ were fixed; the total-effect index *S*_*T*_*i* reflects the overall contribution of *X*_*i*_ to the model’s variance, including its interactions with other parameters.

Numerically, these indices are calculated using the Monte Carlo technique. To ensure effective and uniform coverage of the parameter space, Sobol’ sequences are typically used. A detailed description of the numerical procedure can be found in [6]. In our analysis, the Python package SALib was used [28], [29], where the Sobol’ indices were implemented based on the paper [6]. The number of parameter sets needed for the indices evaluation equals *A* · *N* · (*K* + 2), where A — number of runs per each scenario (1 in most of the experiments), K — number of input parameters of the model (17 or 9 in our experiments for full and truncated sets of parameters respectively), N — number of points in parametric space (should be chosen so that there are enough points for Monte-Carlo integration to converge).

The sensitivity analysis was performed in several steps. First, parameters were sampled and synthetic populations were generated. Then, the spread of the virus was simulated using Covasim. Sobol’ sensitivity analysis was then applied to the model outputs. Two major outcomes were considered: the cumulative number of cases and the cumulative number of deaths. We also present here results for other outcomes: the maximum number of new cases per day, and the maximum number of severe and critical cases in the population, which characterize the peak workload of the healthcare system.

## Results

### Accounting for stochasticity

By definition, Sobol’ sensitivity analysis assumes that the system under investigation is deterministic. However, Covasim, like many other agent-based models, is a stochastic model: identical input parameters may produce different outputs depending on the random seed.

Investigation of the stochasticity effects is presented in Supplementary file 2. Based on the results of the experiment, it was concluded that further analysis should be conducted with a population size of at least 100,000 and 30 initially infected individuals. We chose to use a single run per scenario and to justify this simplification in a separate experiment by explicitly comparing two setups — with and without averaging.

### Sensitivity analysis with full and reduced parameter sets

Sensitivity analysis was initially performed with respect to all 17 input parameters (Supplementary Figure 1). Many parameters had only a minor impact on the variation of the model outputs. To simplify the analysis, eight most ‘important’ parameters were selected, while the remaining parameters were grouped together. Least important parameters were: matrices of community and school contacts, the average number of teacher-to-teacher contacts per person per day, the average number of students per staff member at school, the average number of contacts for non-teaching staff per person per day, average community contacts per person per day, enrollment and employment rate distributions, and school size distribution. From this point on, these parameters were treated as a single combined group in the sensitivity analysis (Fig. 2 A, B; Table 2).

**Table 2.** Sobol’ indices for experiment with truncated set of parameters.

| Input/Output | First order indices |  |  |  |  | Total indices |  |  |  |  |
| --- | --- | --- | --- | --- | --- | --- | --- | --- | --- | --- |
|  | cases | deaths | peak | severe | critical | cases | deaths | peak | severe | critical |
| home contacts | 0.070 | 0.056 | 0.056 | 0.367 | 0.136 | 0.090 | 0.080 | 0.092 | 0.419 | 0.193 |
| work contacts | -0.001 | -0.002 | -0.002 | 0.002 | -0.002 | 0.018 | 0.020 | 0.034 | 0.031 | 0.035 |
| household sizes | 0.720 | 0.192 | 0.624 | 0.004 | 0.052 | 0.748 | 0.344 | 0.665 | 0.122 | 0.208 |
| household head's age by household size | 0.001 | 0.207 | -0.004 | 0.211 | 0.262 | 0.020 | 0.300 | 0.035 | 0.297 | 0.381 |
| students to teachers ratio at schools | 0.010 | 0.000 | 0.012 | 0.004 | 0.003 | 0.039 | 0.019 | 0.060 | 0.040 | 0.039 |
| class size at schools | 0.049 | -0.001 | 0.057 | 0.014 | 0.003 | 0.079 | 0.018 | 0.104 | 0.047 | 0.042 |
| age distribution | 0.002 | 0.335 | 0.001 | 0.187 | 0.307 | 0.022 | 0.516 | 0.036 | 0.300 | 0.480 |
| workplace sizes | 0.105 | -0.002 | 0.185 | 0.073 | 0.018 | 0.125 | 0.020 | 0.215 | 0.102 | 0.056 |
| other parameters | 0.008 | -0.005 | -0.005 | 0.007 | 0.002 | 0.026 | 0.019 | 0.046 | 0.035 | 0.036 |

**Fig 2.**
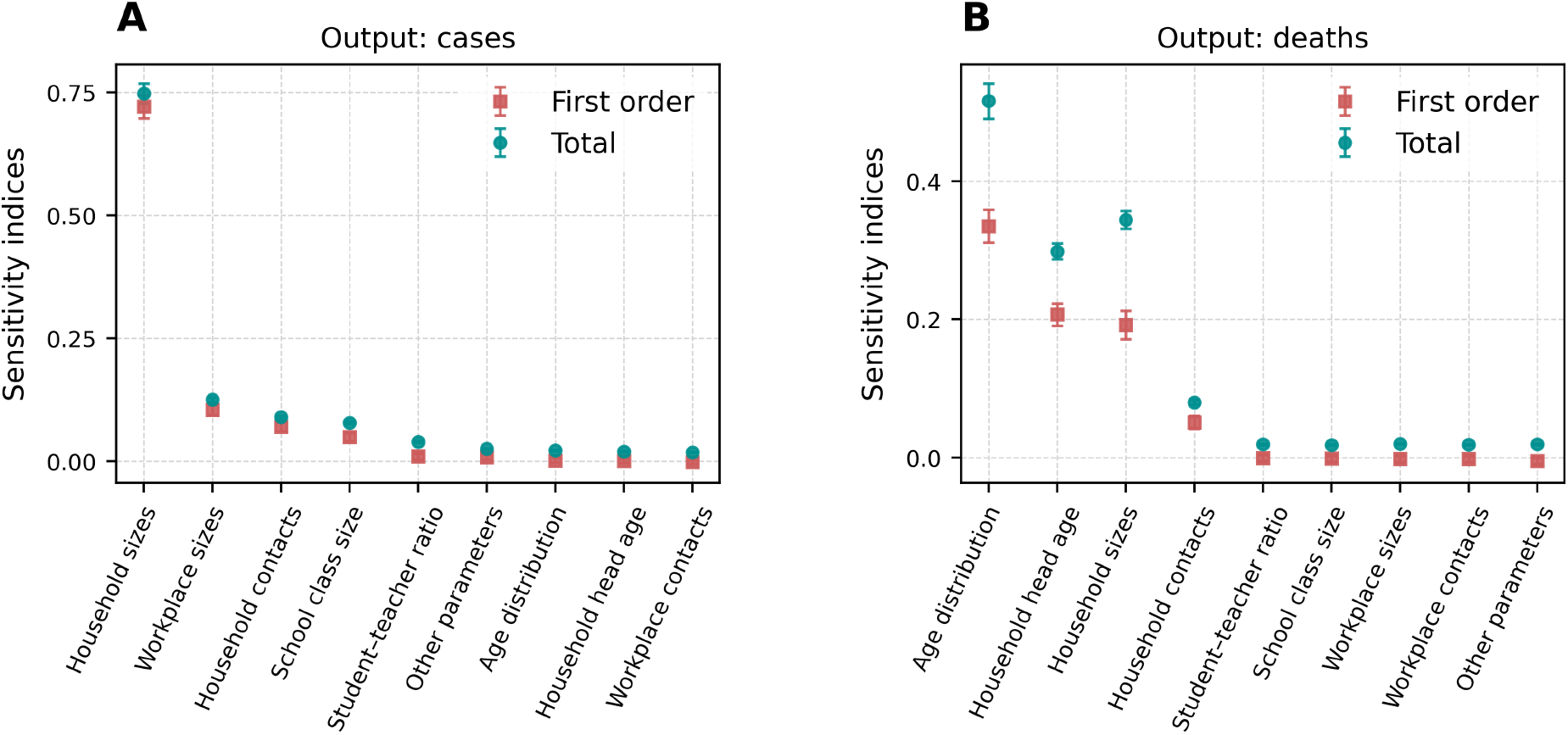
Results of the sensitivity analysis for the truncated set of parameters. (A, B) Sobol’ indices for the cumulative number of cases and deaths, respectively. Red indicates first order Sobol’ indices (S1), while green represents total indices (ST).

As shown in Table 2, the key demographic parameters are the household contact matrix, the distribution of household sizes, the distribution of household head age by household size, the age distribution, and the distribution of workplace sizes.

To ensure that the number of samples in the parameter space was sufficient for the convergence of the Monte Carlo integration, the dependence of Sobol’ indices on the number of samples was examined(Fig. 3 A, B, C, D). Since the indices showed little variation between 1024 and 8192 points, it was concluded that the analysis had converged and was therefore reliable.

**Fig 3.**
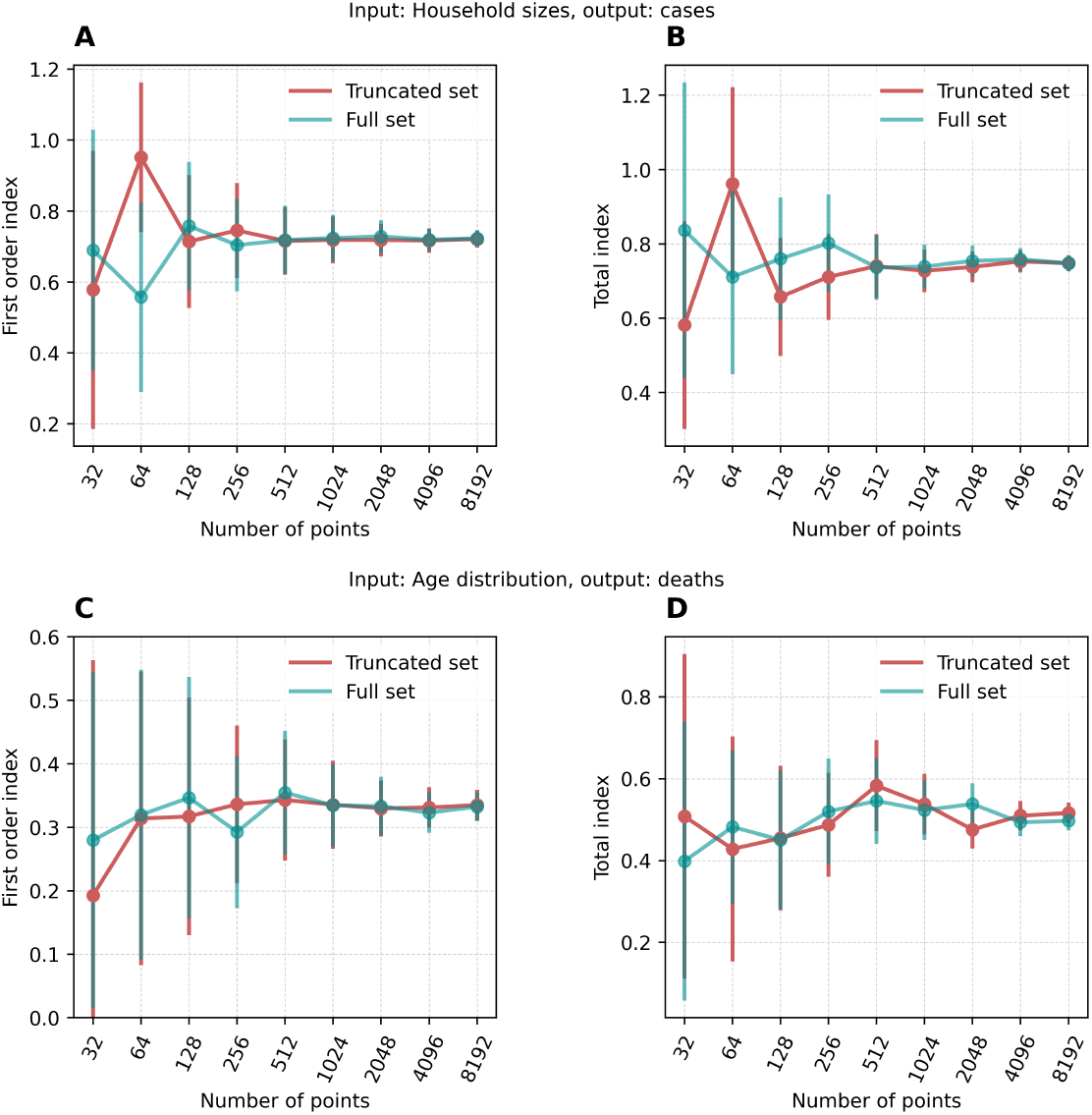
Convergence of Sobol’ indices. Sensitivity indices for the most important parameters with respect to cumulative number of cases (A, B) and deaths (C, D) respectively, shown as a function of the number of sampling points in the parameter space.

To further validate the methodology, we compared the results of the sensitivity analysis with and without averaging over runs. Both analyses were conducted using 1,024 sampling points in the parameter space. The values of the Sobol’ indices did not differ significantly between the two approaches (Supplementary Figure 2 and Supplementary Figure 3), supporting the validity of using a single run per scenario in the main analysis.

As the household size distribution exhibited a major impact on the cumulative number of cases, its role was studied in detail, including the changes caused by altering the algorithm used for household construction in Synthpops. The details can be found in Supplementary file 3.

### Sensitivity analysis for different viruses

The default transmissibility in Covasim (*β* = 0.016) is calibrated to approximate the spread of the original Wuhan strain. To broaden the applicability of our sensitivity analysis to other coronavirus variants and potentially to different pathogens, we repeated the analysis using two additional transmissibility values: *β* = 0.008 and *β* = 0.024 (Fig. 4C and Fig. 4D).

**Fig 4.**
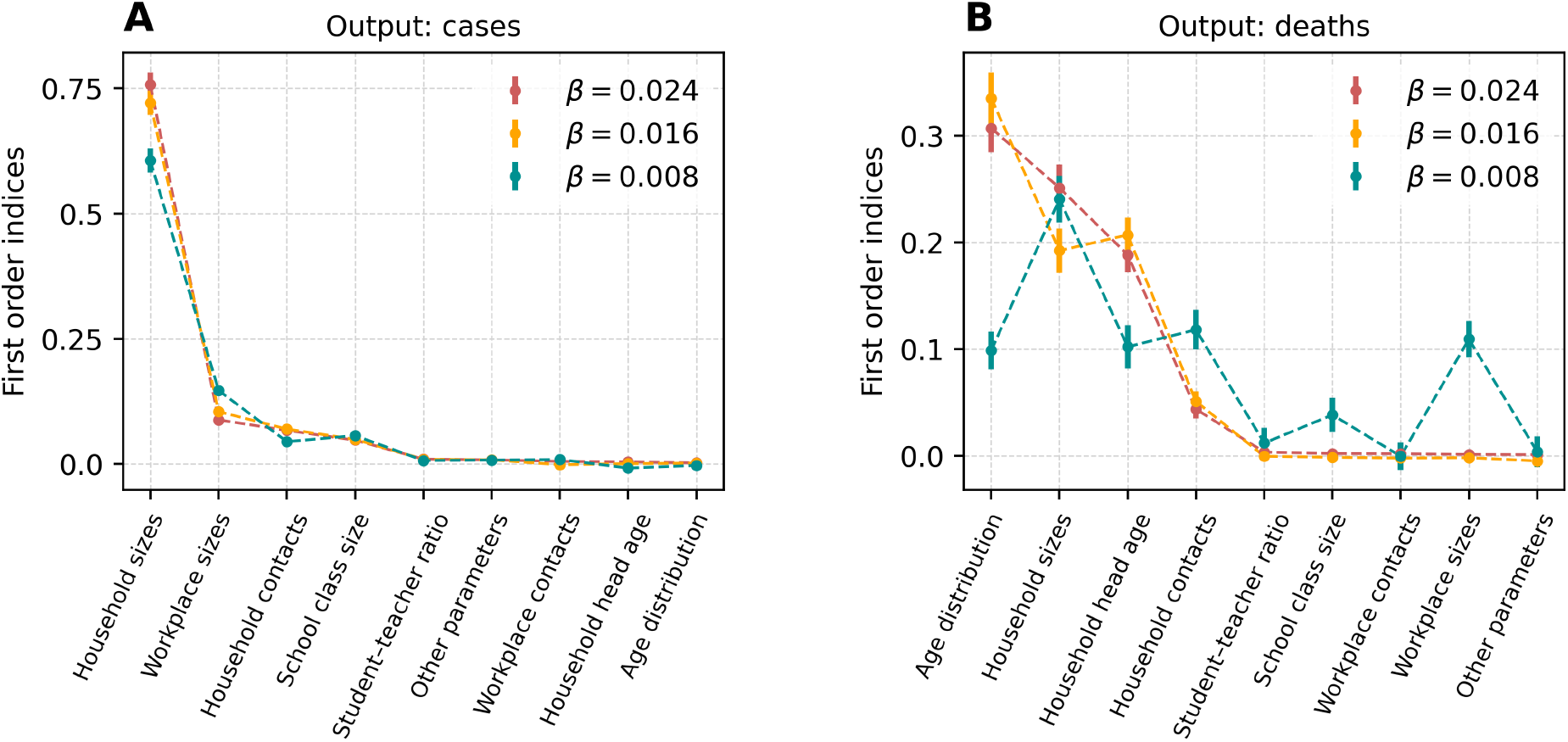
Results of the sensitivity analysis with different virus transmissibility levels. A, B – first order Sobol’ indices for the cumulative number of cases and deaths, respectively, for different values of virus transmissibility; red – 0.024, orange – 0.016, green – 0.008.

The results for the cumulative number of cases were largely independent of the virus transmissibility. For the cumulative number of deaths, the results for *β* = 0.016 and *β* = 0.024 were closely aligned, while those for *β* = 0.008 differ significantly. This distinct behavior at low transmissibility can be explained by epidemic saturation in the other two scenarios: for *β* = 0.024 (and in almost all cases for *β* = 0.016), the number of infections exceeded the total population size (10^5^), indicating that nearly all individuals were infected at least once (Supplementary Figure 4 A). Consequently, the cumulative number of deaths was similarly distributed for these two values (Supplementary Figure 4 B). In contrast, in the case of low transmissibility (*β* = 0.008), the number of cases remained below the population size in all simulations, and the number of deaths was substantially lower. As a result, the sensitivity of the outcomes to individual parameters changed under this scenario.

## Discussion

Identifying the parameters that significantly influence model outcomes is an important step in computational epidemiological simulations. Parameters of low importance can be inferred with less precision, whereas parameters with high impact must be accurately estimated and regularly updated to ensure model reliability. By using autoencoders, we were able to analyze the Covasim model with Sobol’ indices and to investigate the importance of non-scalar population parameters, which inform parameter prioritization.

Our results highlight that only a subset of demographic parameters exerts a significant influence on the model output. In particular, the household size distribution, age distribution, household contact matrix, matrix of household head age by household size, and workplace size distribution influenced both the total number of infections and the number of deaths. These findings are consistent across multiple experimental settings, including variations in virus transmissibility and different synthetic population generation methods. Notably, all of the most important parameters were vectors or matrices. This underscores the importance of the approach proposed in this study for analyzing non-scalar parameters.

Many of the important parameters were related to household characteristics. This can be explained by the higher transmission efficiency of contacts within households compared to other contact layers [37]. Moreover, household features—such as age composition and size distribution—vary substantially across countries, regions, and cities, reflecting differences in socioeconomic and cultural contexts. Since these parameters exhibit substantial variability in real-world data, sampling from the latent space of autoencoders trained on such data resulted in a wide range of realistic values, thereby increasing the overall importance of these parameters in the model.

The Sobol’ method of sensitivity analysis, which we used to rank the parameters by their importance, is convenient due to its straightforward implementation and the ease of interpreting the results. However, it does not account for stochasticity. To reduce stochastic effects, we used a relatively large initial number of infected individuals (30). In this setup, the difference between runs with identical inputs was a few percent in both the number of cases and deaths. When thousands of runs with shuffled parameter values were used to evaluate the Sobol’ indices, the effect of model stochasticity was negligible (Supplementary Figure 2, Supplementary Figure 3). There are some limitations to the methods and results of our study.

Our sampling method assumed the input parameters to be independent, as it is a necessary condition for clear interpretation of a variance-based sensitivity analysis [6]. For real populations, this is not the case: for example, the dependence of the household head age on the household size cannot be independent of household size distribution and age structure of the population. As a result, in some cases, SynthPops was unable to construct a valid synthetic population from certain parameter combinations, and these samples had to be excluded from the analysis. In future work, this limitation can be addressed by training a single autoencoder to jointly reconstruct all demographic parameters, allowing for the generation of coherent and realistic parameter sets. The theoretical derivation and practical application of Sobol’ indices for models with correlated inputs can be found in [38].

Sampling via autoencoders benefits from the simplicity of the approach and the models’ ability to capture structural patterns in demographic parameters. Nevertheless, some information is inevitably lost in their low-dimensional representations. This study employed relatively simple architectures, leaving room for future improvements through the use of more sophisticated models.

Inevitably, the results of the sensitivity analysis depend on the method of parameter sampling, including the bounds in which parameters vary. In our study, we determined these bounds based on real-life data where enough data was available. In other cases, we used the default parameter values from Covasim and applied a scaling factor selected from [0.5, 2] range. Broader ranges led to frequent failures in the construction of synthetic populations.

Lastly, we didn’t vary some parameters which interact with demographic parameters and thus influence the results of the sensitivity analysis. For example, in the default settings of the Covasim contacts within the households are ten times more likely to result in infection than contacts within the workplaces (relative transmissibility of the virus is multiplied by 3 for household contacts and by 0.3 for workplace contacts). This leads to the increased sensitivity of the outputs to the household parameters in comparison with sensitivity to the workplace parameters, but this disposition could have changed if we had included these Covasim coefficients in the analysis. This constrains our estimates to a epidemiological profile of the virus.

## Conclusion

In this study, we presented a novel autoencoder-based sampling method for multidimensional parameters and conducted a global sensitivity analysis of the agent-based epidemiological model Covasim, focusing on the demographic parameters used to construct synthetic populations. By applying Sobol’ sensitivity indices we evaluated the relative importance of scalar, vector, and matrix parameters on key epidemiological outcomes such as cumulative cases and deaths.

Our results highlight that only a subset of demographic inputs —– particularly the household size distribution, age distribution, household contact matrix, matrix of household head age by household size, and workplace size distribution —– exert a dominant influence on the chosen model outputs. These findings are consistent across multiple experimental settings, including variations in virus transmissibility and different synthetic population generation methods. Due to the dominant influence of non-scalar parameters on the outputs of the model

Our method can be useful for sampling realistic, data-driven parameters and is therefore particularly well-suited for researchers employing agent-based models.

## Data Availability

All code used for this study is available online at https://github.com/tsurkisvera/sensitivity-analysis-covasim

https://desapublications.un.org/publications/world-population-prospects-2024-summary-results

https://zenodo.org/records/8142652

https://data.un.org/Data.aspx?d=POP&f=tableCode%3A50

https://www.oecd.org/en/publications/education-at-a-glance-2024_c00cad36-en.html

https://rshiny.ilo.org/dataexplorer18/?lang=en&id=EMP_TEMP_SEX_AGE_NB_A

https://ec.europa.eu/eurostat/databrowser/view/SBS_SC_SCA_R2__custom_2928090/bookmark/table?lang=en&bookmarkId=4bdbd2d1-3236-4d2f-be66-c77585a6619e

## Supplementary data

Supplementary data are available at *MedRxiv* online

## Data availability

The code used for the experiments is publicly available at https://github.com/tsurkisvera/sensitivity-analysis-covasim.

