## Supplementary figures and images for "Demographic Drivers of Epidemic Outcomes: Sensitivity Analysis of Multidimensional Parameters in the Covasim Model"

### Supplementary Figure 1

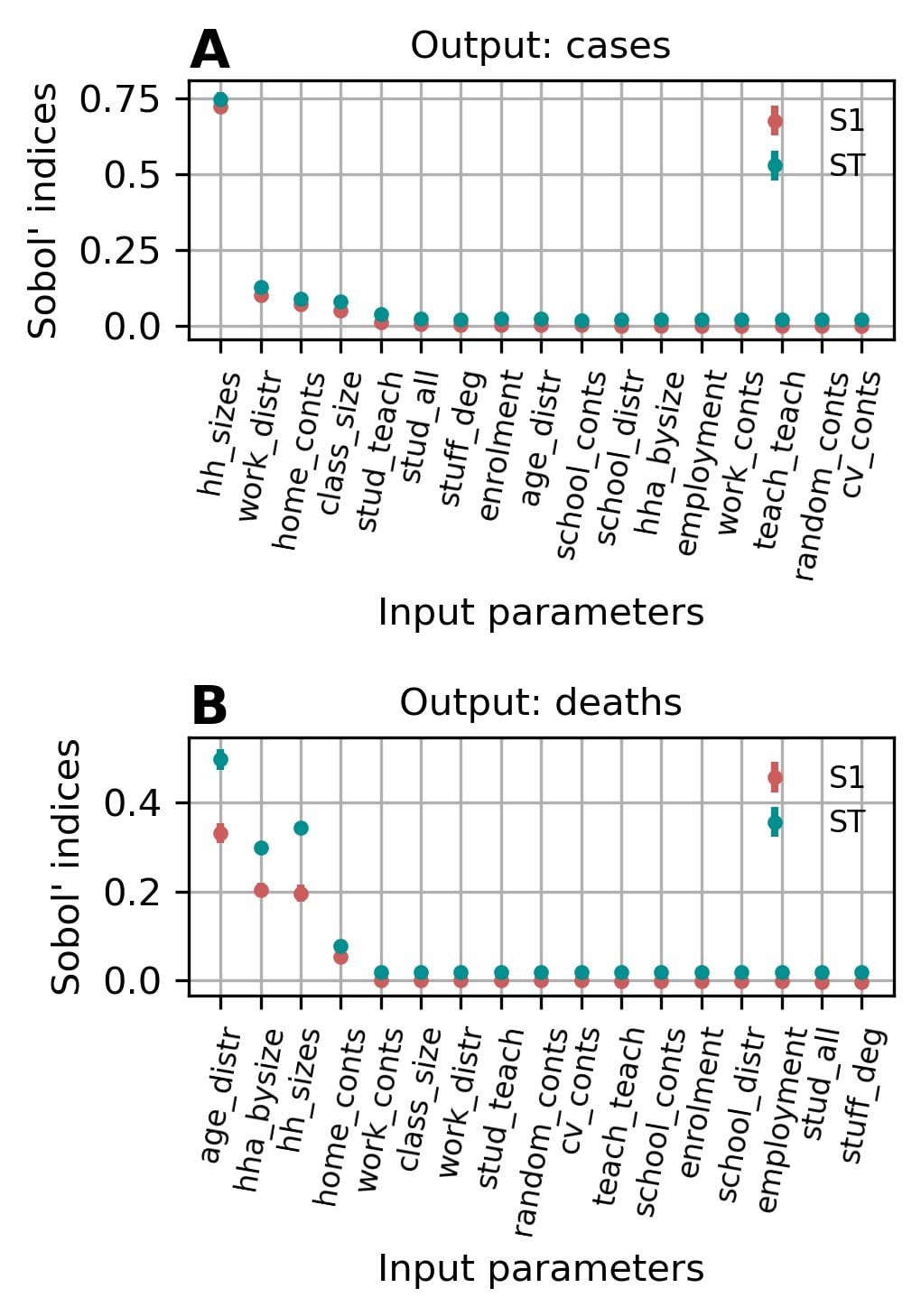

### Supplementary Figure 2

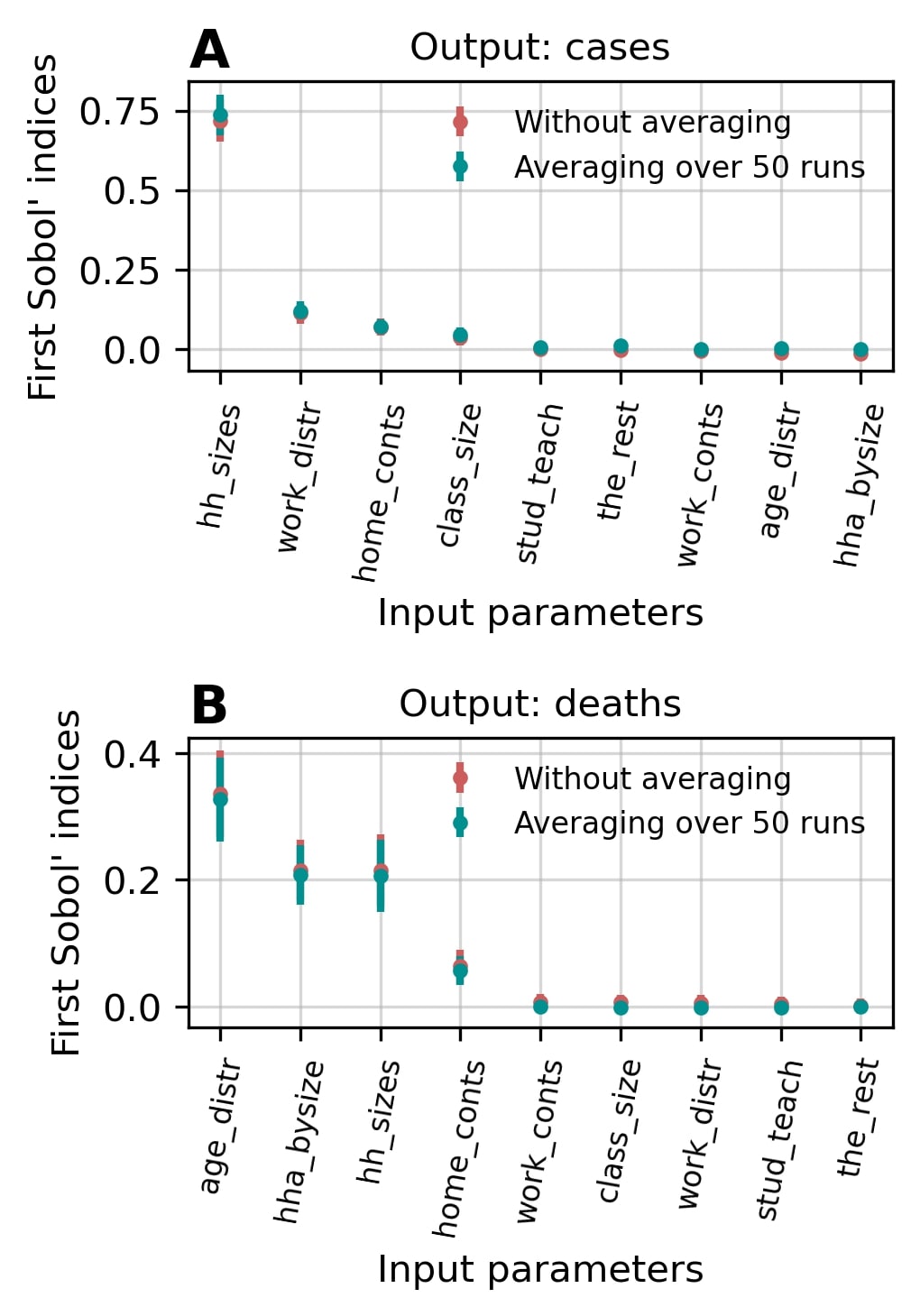

### Supplementary Figure 3

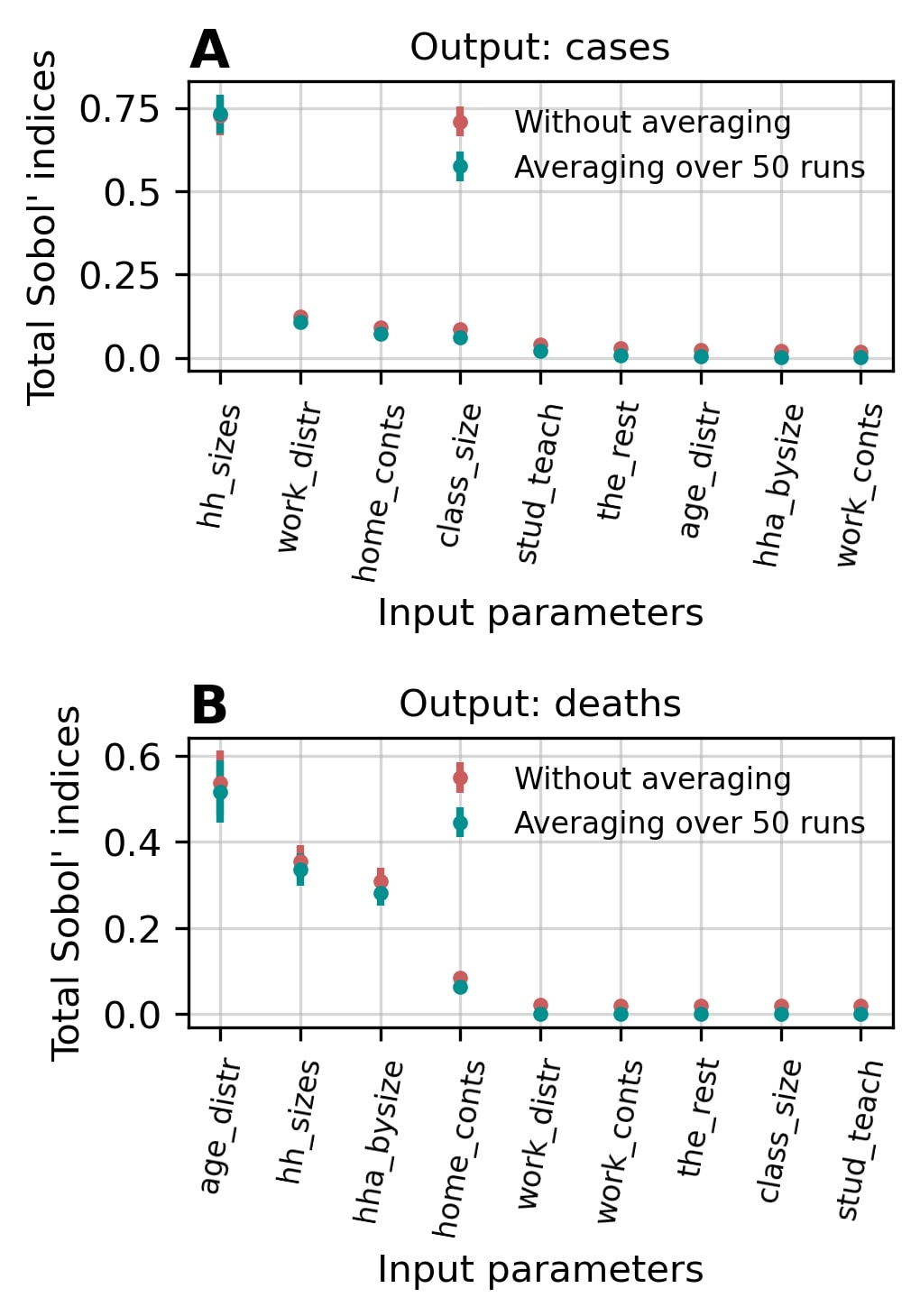

### Supplementary Figure 4

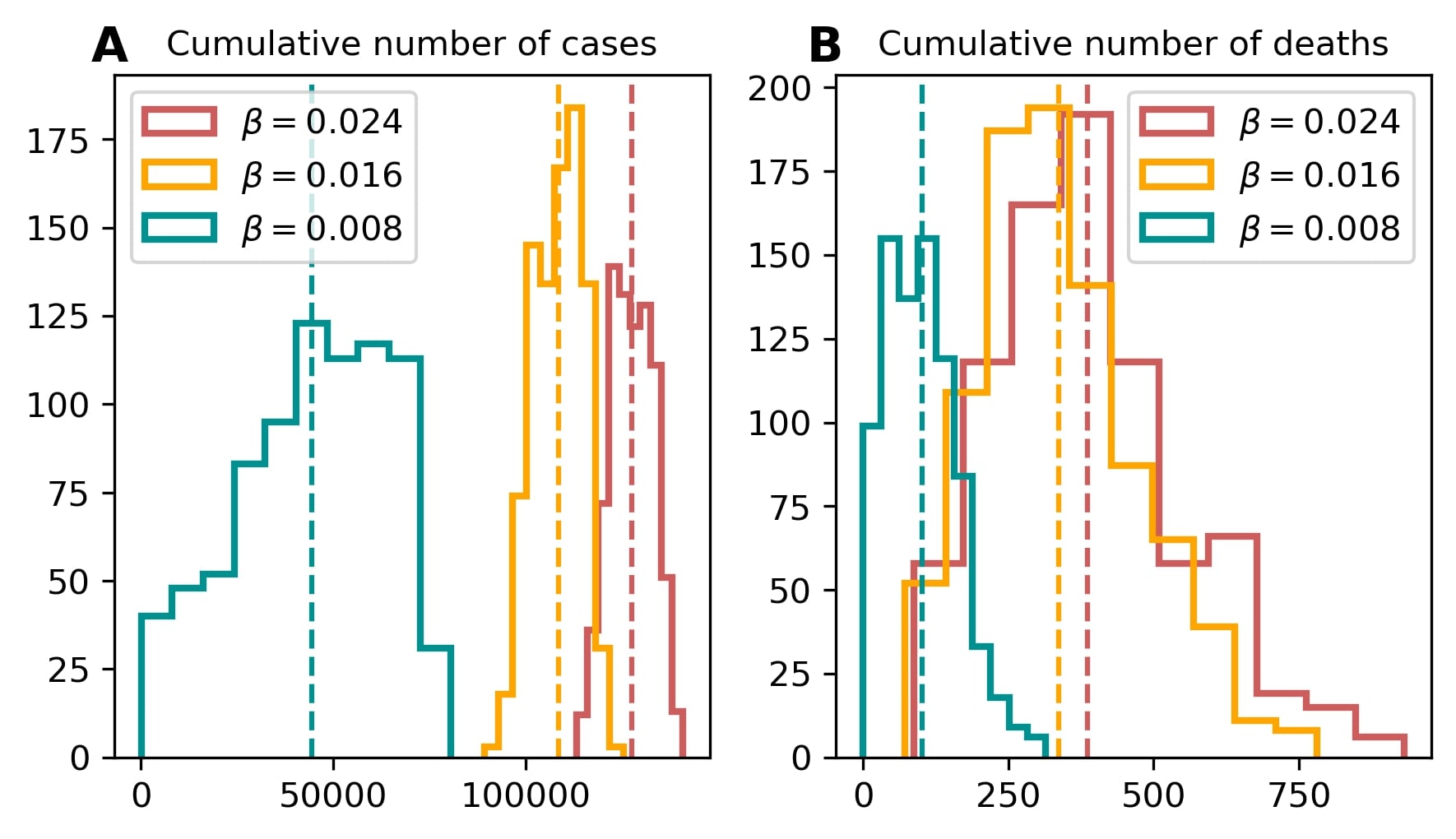
