## Supplementary File 3 for "Demographic Drivers of Epidemic Outcomes: Sensitivity Analysis of Multidimensional Parameters in the Covasim Model"

### Cumulative number of cases: Are household sizes really so important?

The distribution of household sizes had a major impact on the cumulative number of cases $S1 = 0.7$. To further investigate this effect, the outputs of Covasim under varying household size distributions were explored, while keeping all other parameters fixed at their default values.

The latent space of the autoencoder trained on household size distributions was divided into 100 segments. For each segment, a corresponding household size distribution was generated, and 200 simulations were performed per scenario. A similar experiment was conducted for the household contact matrix as the only varied parameter.

The distribution of household sizes had indeed more influence than the household contact matrix for both cumulative case counts and cumulative deaths (Fig. 1 A, B). Furthermore, the top panel of Fig. 1 shows that the number of cases increased with a higher proportion of large households (5–7 members), whereas the number of deaths tended to decrease. The epidemic curves for the selected points in the latent space of the household size distribution autoencoder supported the increase of the cumulative case counts with the increase of the fraction of large households (Fig. 1 E, F).

As the observed relationship between the number of cases and deaths contradicted common expectations, an alternative setup was explored. In SynthPops, there are two methods for household generation. In all previous experiments, the default method was used. However, the developers note in the code that for arbitrary populations the alternative method is more appropriate [1]. Following this recommendation, the analysis was repeated using the alternative method by setting the parameter `household_method` to `fixed_ages` instead of `infer_ages` (Fig. 1 C, D).

With the alternative household construction method, the number of deaths behaved consistently with the number of cases, suggesting a more coherent epidemic response pattern. Despite this, the default method was retained in subsequent experiments, as it provided greater flexibility across the entire parameter space. In contrast, the stricter `fixed_ages` method frequently failed to generate valid populations for many parameter combinations. Specifically, in the truncated parameter set analysis using 8,192 sampling points, a valid population was constructed for only about one third of the parameter sets, resulting in 6% successful evaluations, because to include the point in the sensitivity analysis all auxiliary populations should have been constructed as well [2].

Despite frequent failure to generate populations, the analysis using the alternative household construction method still converged, and the resulting Sobol' indices were compared with those from the main experiment (Fig. 2, Fig. 3).

Despite the reduced number of successful runs and differences in population construction, the computed sensitivity indices remained consistent, supporting the robustness of the overall analysis.

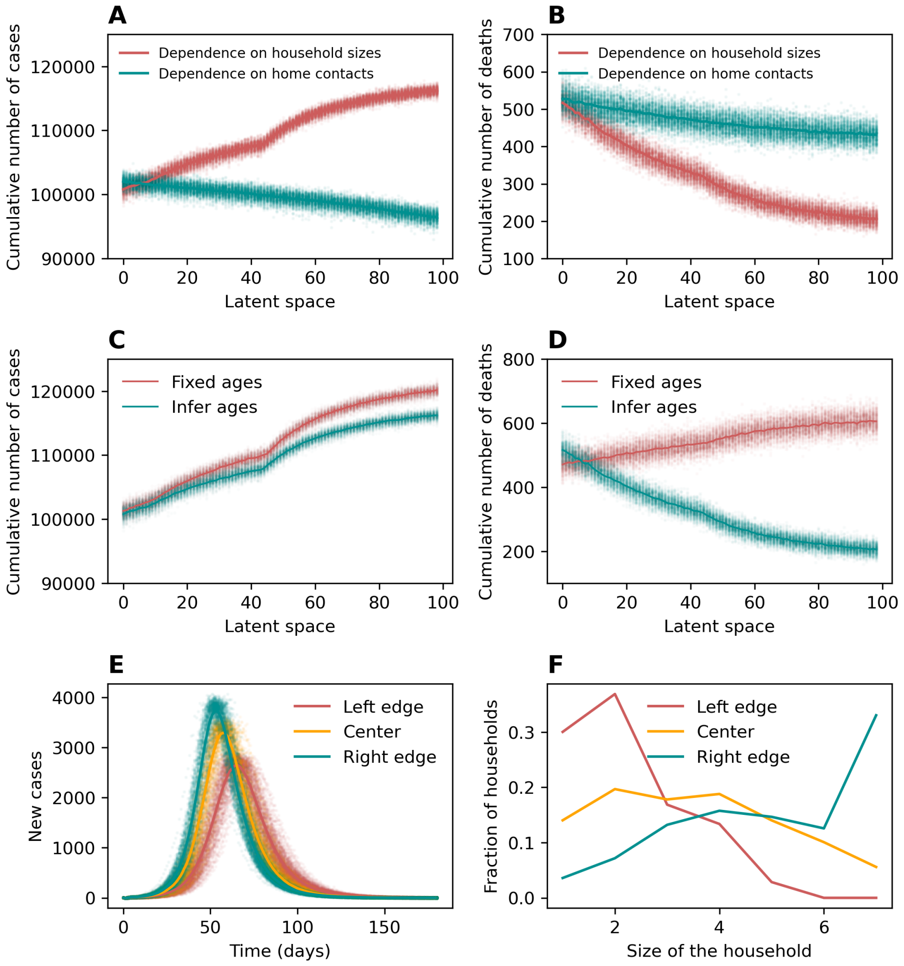


Figure 1: **Exploring the effect of household size distributions.** (A, B) Dependence of cumulative number of cases and deaths respectively on the position in the latent space that defines the household size distribution (red) and the household contact matrix (green). The horizontal axes indicate indices of points uniformly sampled from the one-dimensional latent spaces of the corresponding autoencoders. (C, D) Comparison of the behavior of the cumulative number of cases and deaths respectively for two different household construction methods: the default method (`infer_ages`, green) and the alternative method (`fixed_ages`, red). (E) Epidemic curves for three selected points in the latent space of the household size distribution autoencoder (red – left edge, orange – center, green – right edge of the segment). (F) Corresponding household size distributions; colors match those in panel E.

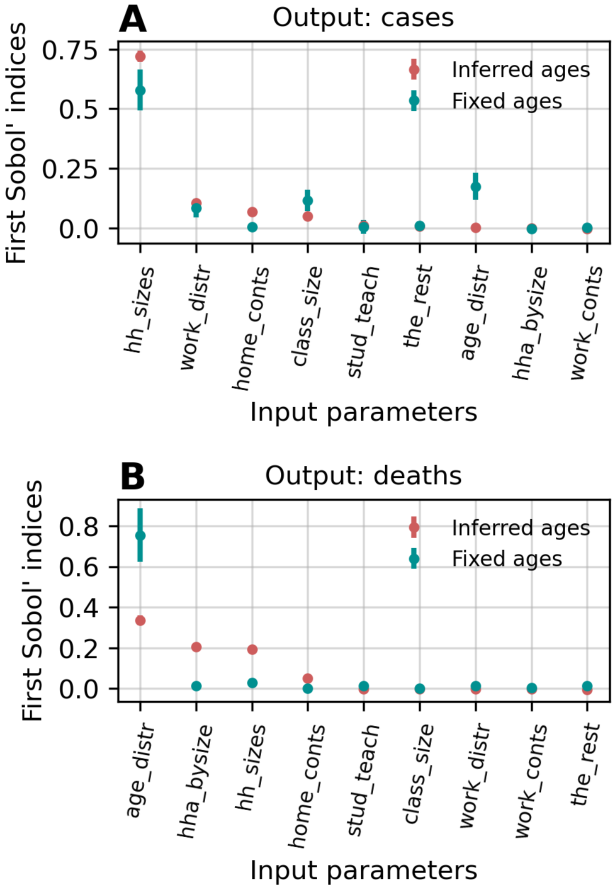


Figure 2: **Results of the sensitivity analysis with the alternative method of household construction: first order indices.** First order Sobol' indices are shown for the cumulative number of cases (A) and deaths (B). Analysis was conducted using 8192 points in parameter space. Red points correspond to the sensitivity analysis with the default constructing method ('infer_ages'), green points – to the analysis with the alternative method('fixed_ages').

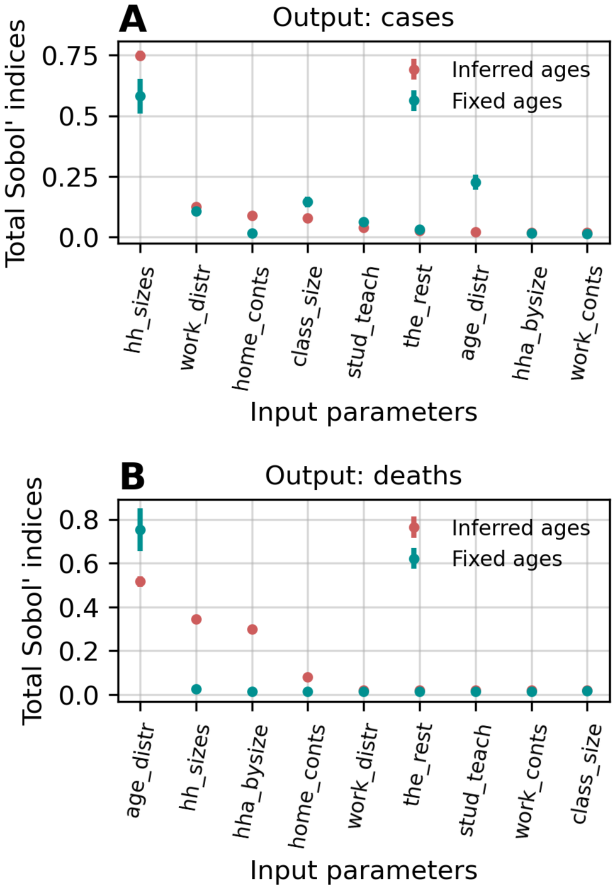


Figure 3: **Results of the sensitivity analysis with the alternative method of household construction: total indices.** Total Sobol' indices are shown for the cumulative number of cases (A) and deaths (B). Analysis was conducted using 8192 points in parameter space. Red points correspond to the sensitivity analysis with the default constructing method ('infer_ages'), green points – to the analysis with the alternative method('fixed_ages').

### Sensitivity analysis without varying household sizes

Given the substantial impact of the household size distribution on simulation outputs, additional experiments were conducted in which this distribution was held constant. Specifically, three fixed household size distributions were selected, each corresponding to a distinct point in the latent space of the respective autoencoder (as shown in Fig. 1F). Sensitivity analysis was then performed separately for each of these fixed distributions.

The results are presented in Fig. 4A and Fig. 4B. It can be observed that as the proportion of large households increases, the importance of the household contact matrix and the matrix of household head age by household size also increases. In contrast, the importance of the overall age distribution and the workplace size distribution decreases.

This observation can be explained as follows: when large households dominate, the majority of infections occur within households, making other contact types less important. In contrast, when smaller households are more common, workplace interactions become more significant in driving the spread of the infection.

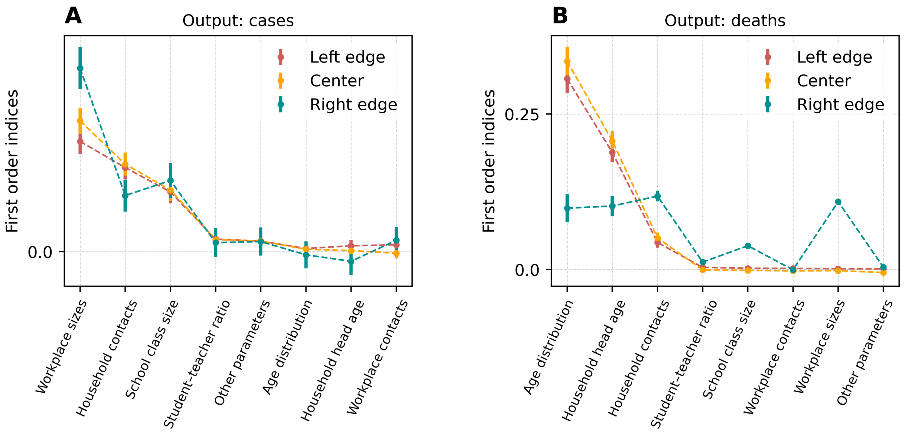


Figure 4: **Results of the sensitivity analysis with different household size distributions.** A, B – first order Sobol' indices for the cumulative number of cases and deaths, respectively, for different household size distributions; colors correspond to those in the bottom panel of Fig 1: red – left edge of the latent space of the autoencoder for household size distribution (small households), orange – center, green – right edge (large households).
