## Supplementary File 2 for "Demographic Drivers of Epidemic Outcomes: Sensitivity Analysis of Multidimensional Parameters in the Covasim Model"

### 1 Accounting for stochasticity

The behavior of the outputs' variance upon averaging over different numbers of simulation runs was examined. For a given number of repetitions (ranging from 1 to 700), ten independent sets of simulations using identical input parameters were performed. Within each set, the outputs from Covasim were averaged, and the variance was then evaluated across the ten averaged values (Fig. 1). It was concluded that further analysis should be conducted with a population size of 100,000 and 30 initially infected individuals.

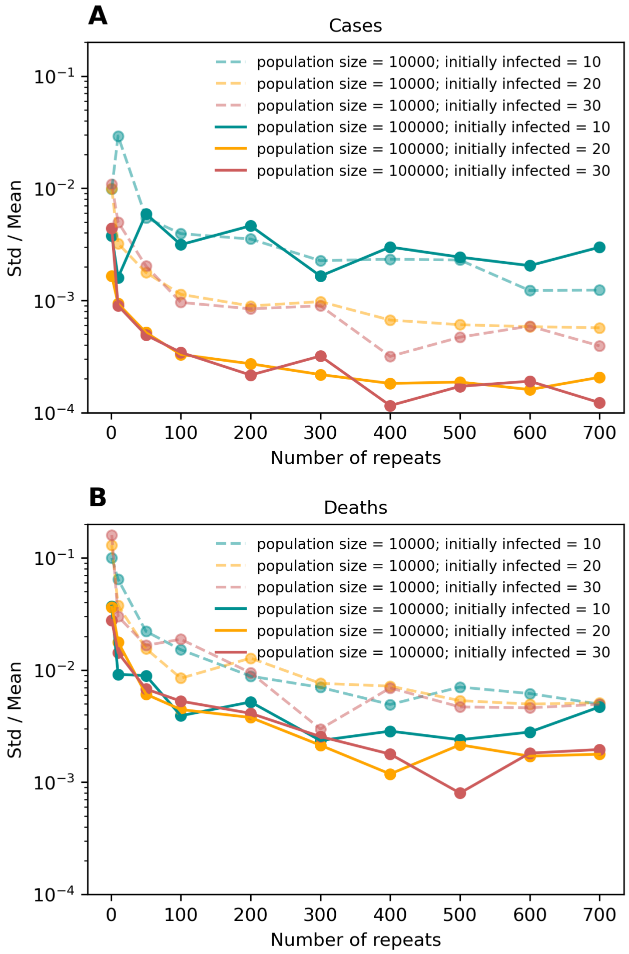


Figure 1: **The behavior of the dispersion in the outputs as a function of the number of runs per scenario.** Ten sets of chosen numbers of scenarios with equal parameters were run on different random seeds. Outputs of these runs were averaged within every set. The ratios of the standard deviation to the mean for different numbers of repeats are shown for the cumulative number of cases during the outbreak (top picture) and for the cumulative number of deaths (bottom picture). We repeated our experiment for different initial numbers of infected people (10 – green color on the plots, 20 – orange color, or 30 – red color) and different sizes of population (10.000 – dashed line or 100.000 – solid line).
