## Supplementary File 1 for "Demographic Drivers of Epidemic Outcomes: Sensitivity Analysis of Multidimensional Parameters in the Covasim Model"

### Learning curves of all autoencoders trained in this work

#### Age distribution

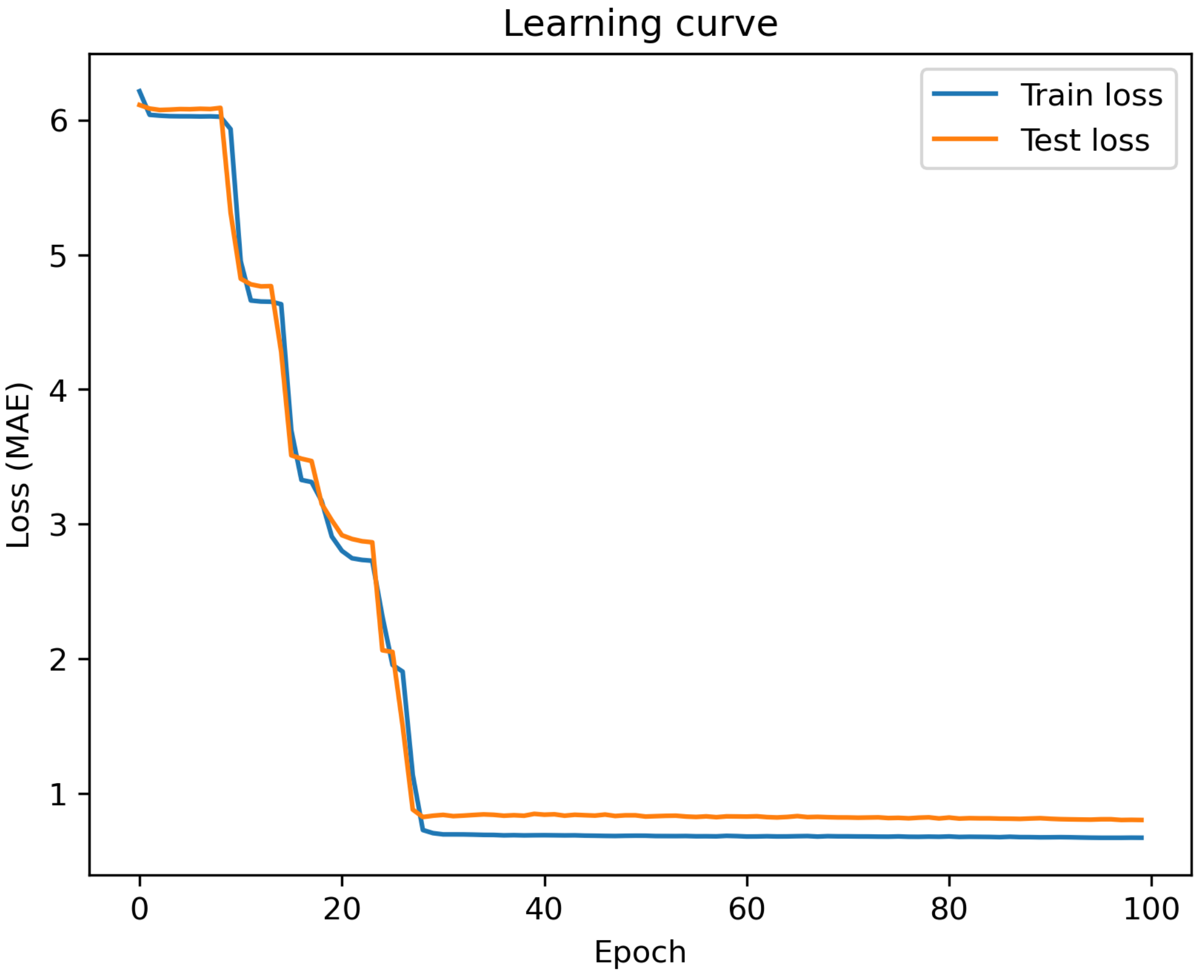

### Household size distribution

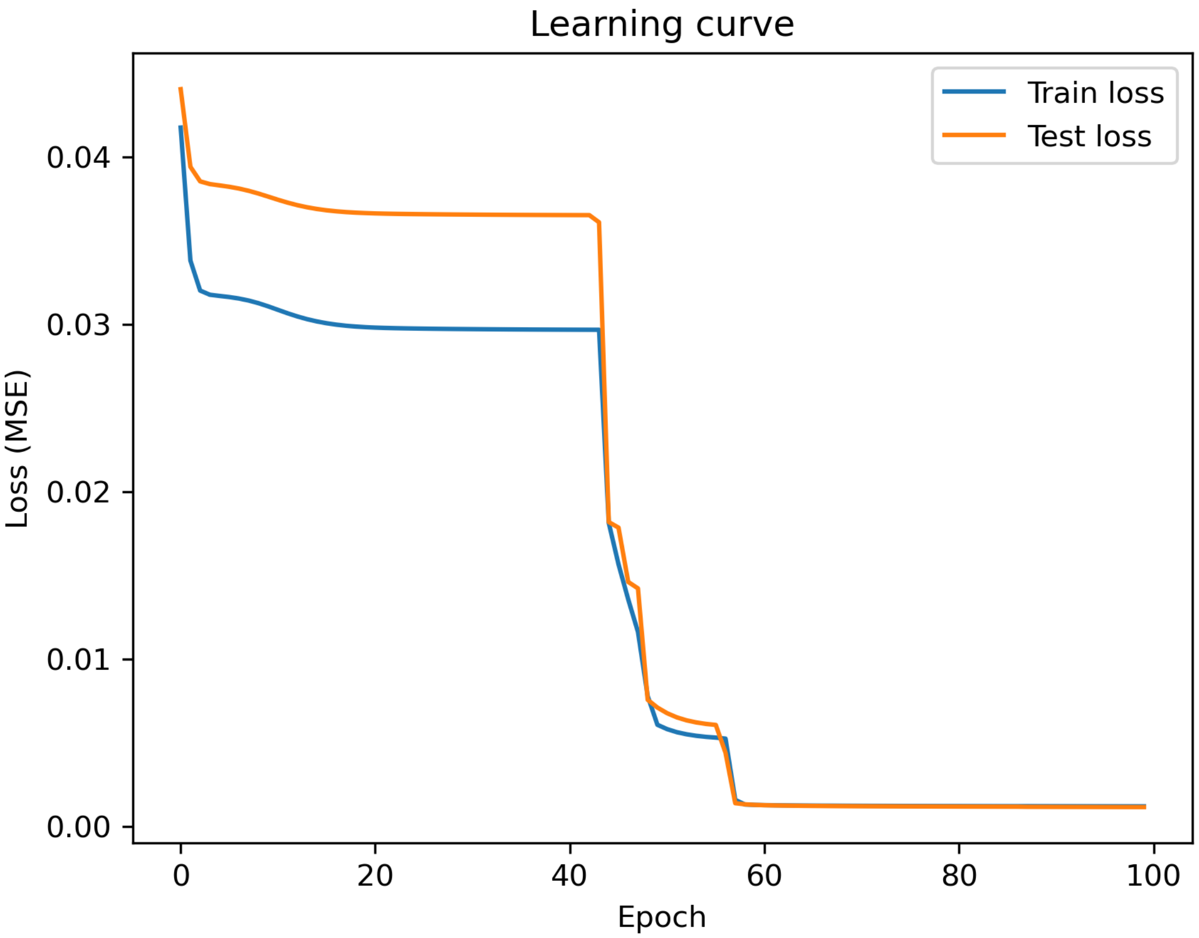

### Household head age by household size

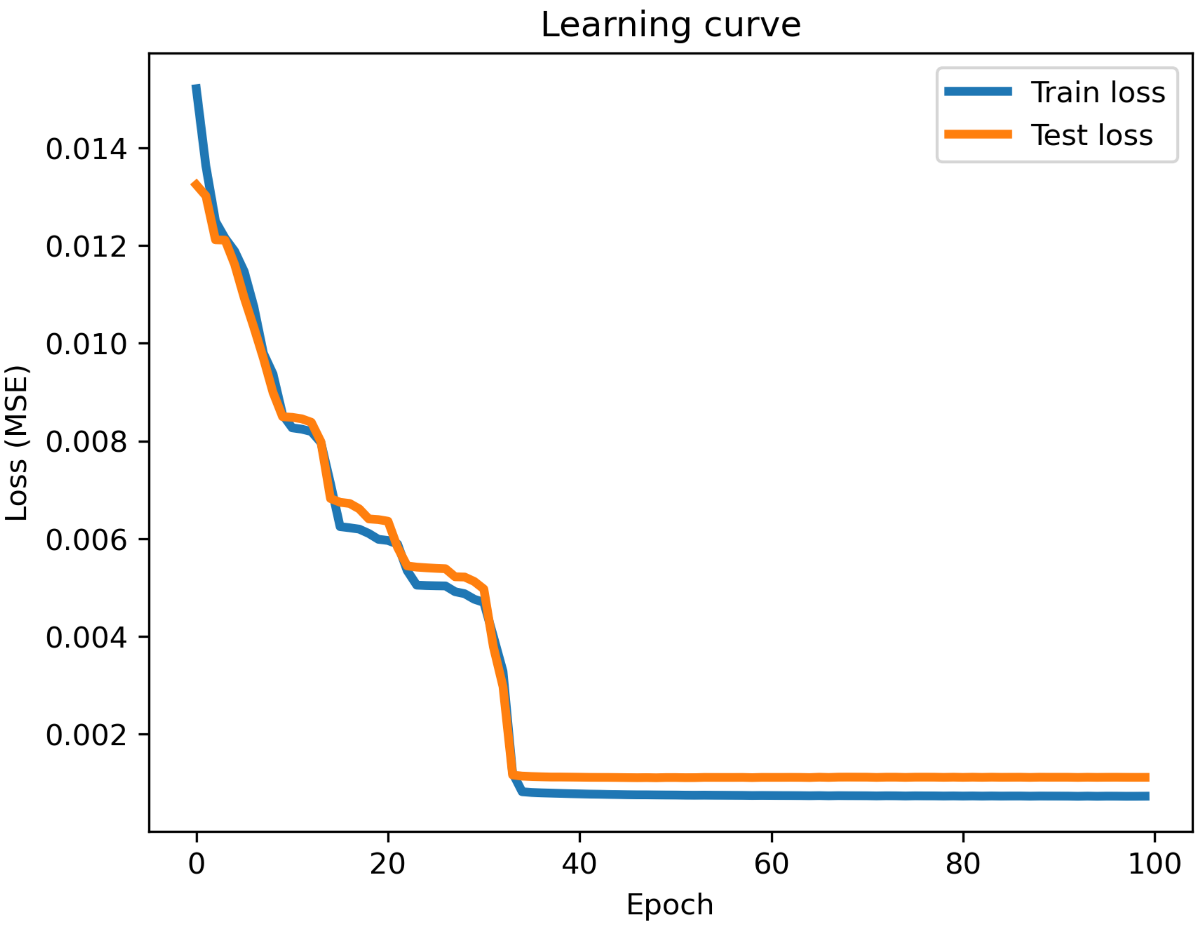

Household contact matrix

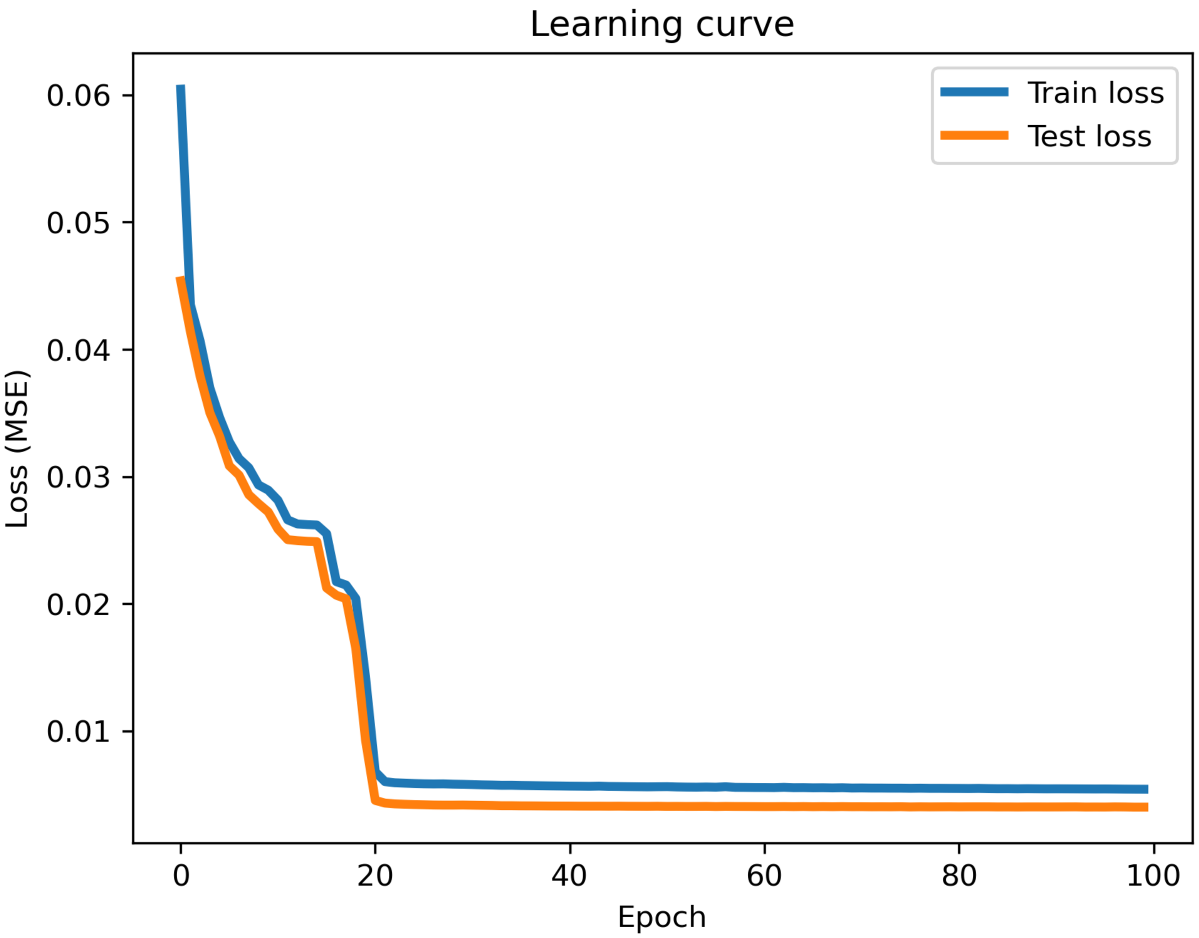

### Workplace contact matrix

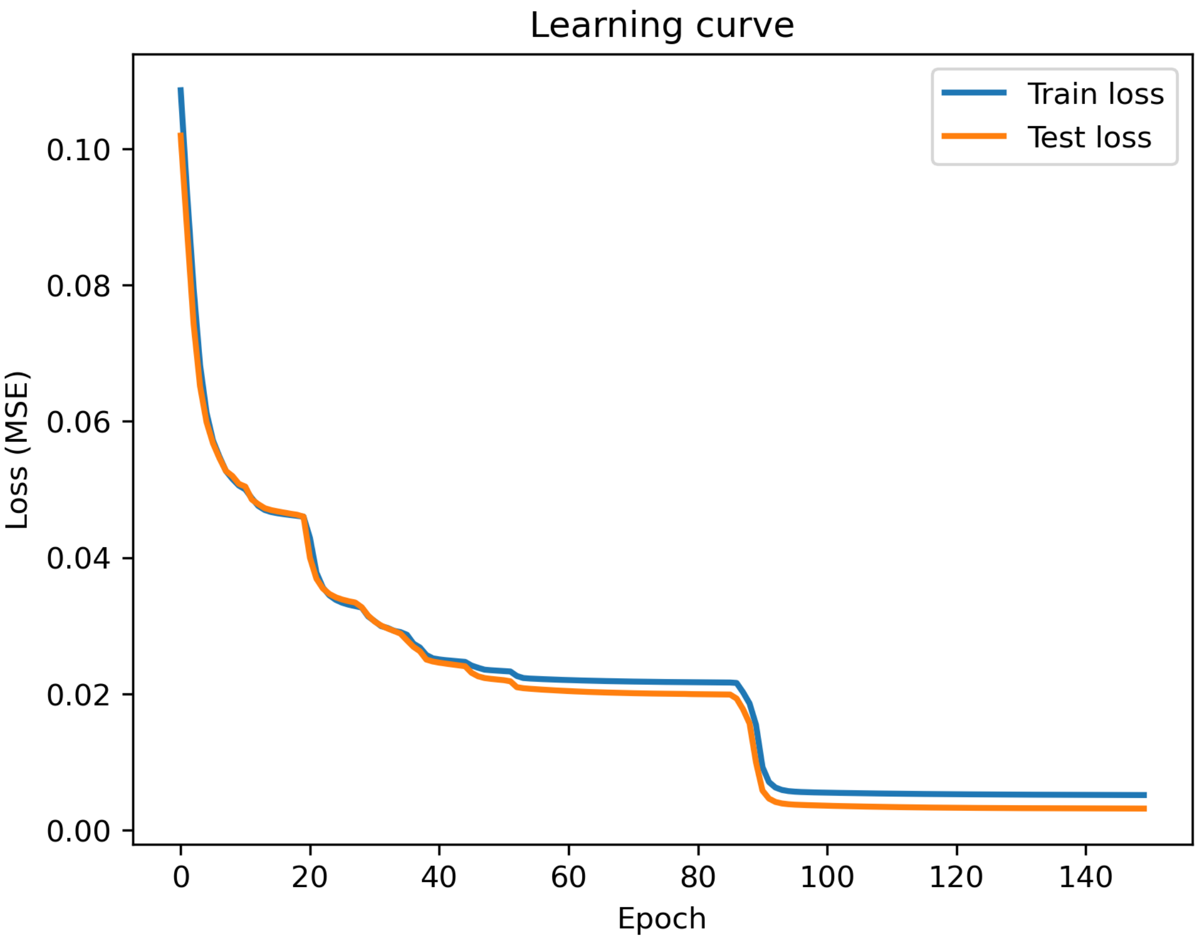

### School contact matrix

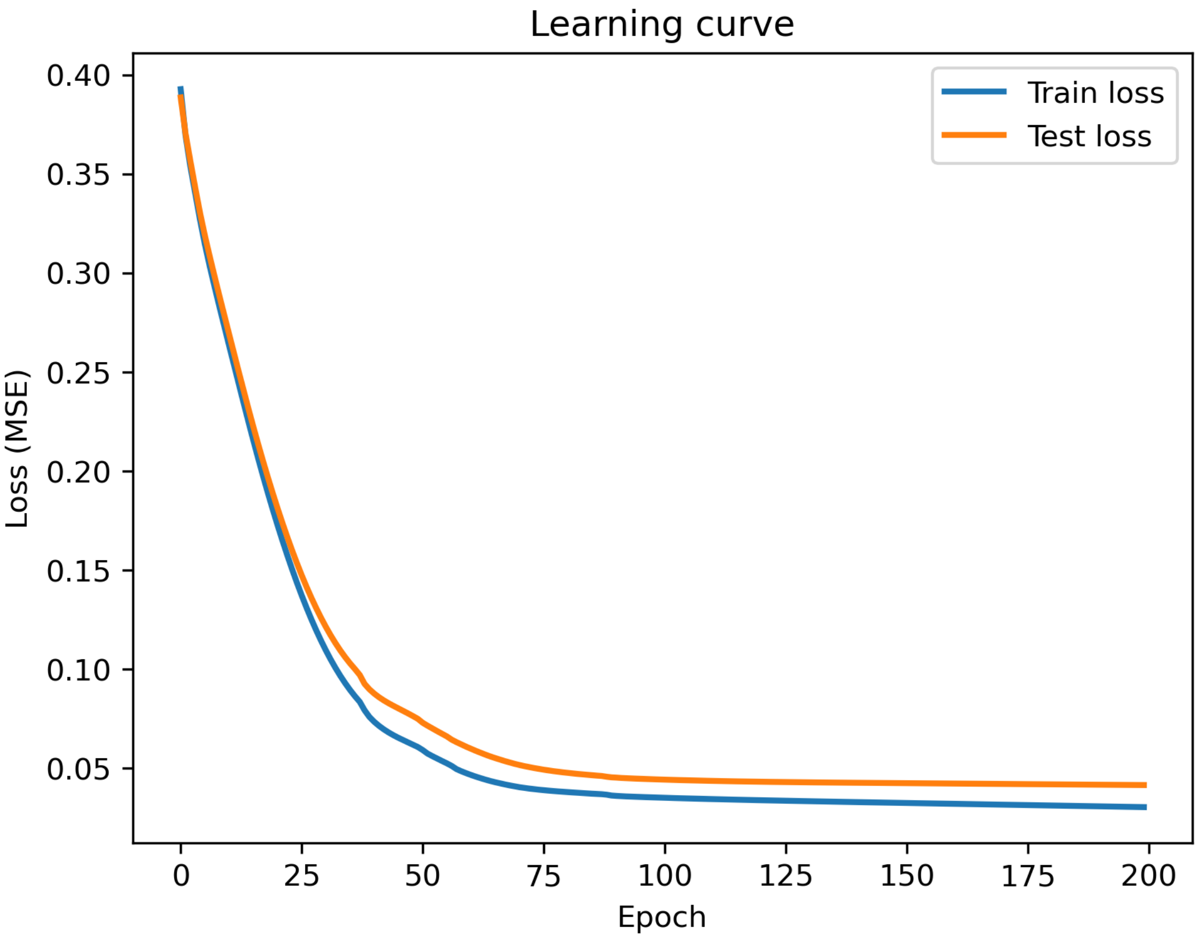

### Random contact matrix

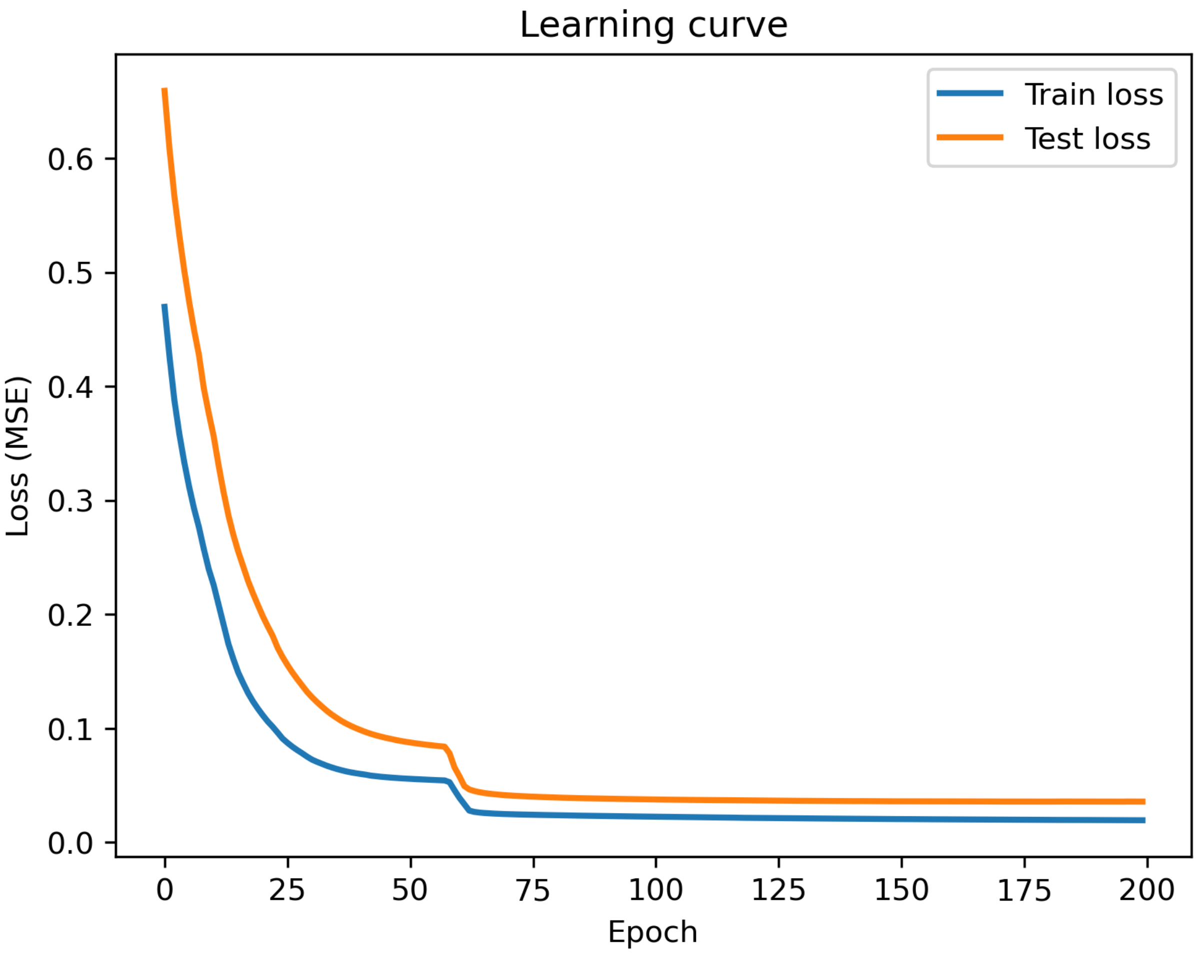

### Employment rate

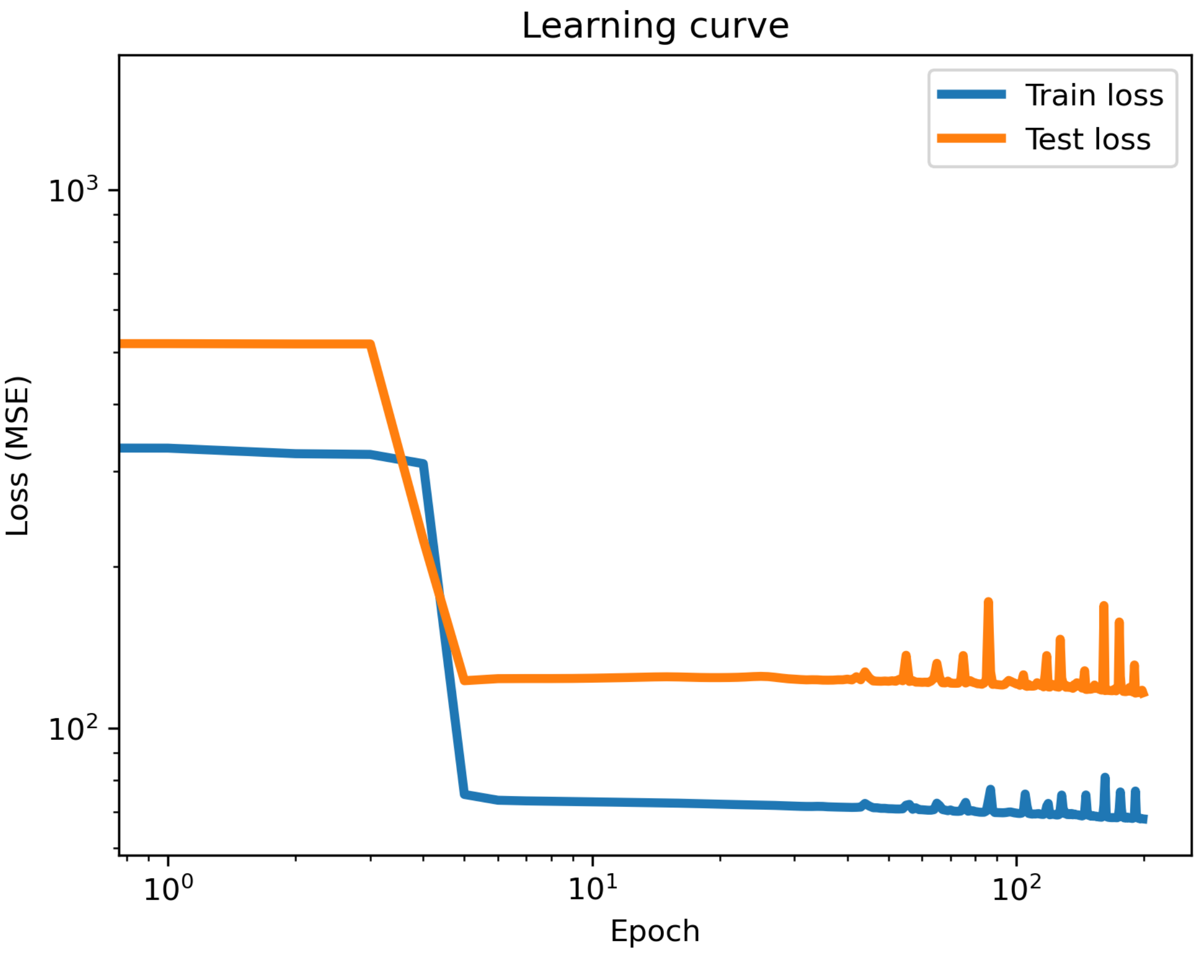
